# Protocol for research examination of individual suicides occurring in chronic pain: A qualitative approach to psychological autopsy methodology

**DOI:** 10.1101/2025.08.01.25332647

**Authors:** Stefan G. Kertesz, Allyson L. Varley, April E. Hoge, Thomas E. Joiner, Beth Darnall, Kate Nicholson, Anne Fuqua, Mary F. Gilmore, Kevin R. Riggs, Stephanie Gamble, Adam J. Gordon, Yogesh Dwivedi, Mark Flower, Christin Veasley, Carla Stumpf Patton, Ashley Leal, Dawn Gibson, Jim Elliott, Megan B McCullough

## Abstract

**Introduction:** In the United States, taper and discontinuation of opioids prescribed for long-term pain have emerged as statistical correlates of suicidal events. Suicide is a complex and multidetermined event reflecting a combination of risks occurring over time in a particular narrative context. Prevention of suicides should be informed by a detailed understanding of life events, pain-related and other risk factors contributing to these tragedies. To date, there have been no efforts to qualitatively profile these suicides through interview of bereaved survivors or review of medical records. This method is usually termed “psychological autopsy.”

**Aims:** This paper summarizes the protocol for the Clinical Context of Suicide Following Opioid Transitions (CSI:OPIOIDs) study. The study seeks to qualitatively characterize patient and clinical context factors associated with suicide among persons who died by suicide in the context of opioid stoppage or reduction, and to compare findings between Veteran and non-Veteran decedents.

**Methods:** In the United States, there is no master list for suicide deaths linked to an antemortem health care event. For this reason, recruitment requires public advertising followed by screening of bereaved individuals who wish to participate. Data collection and interpretation are guided by the Social-Ecological Model for suicide. The study involves a collaboration of persons with lived experience and disciplinary experts in suicide, primary care, pain, health services, and medical anthropology.

**Conclusions:** This study aims to deliver the first in-depth analysis of suicide events occurring in persons with chronic pain who died by suicide in the context of a prescription opioid reduction or stoppage. The results should provide insights that can guide alterations to care by health systems and by individual practitioners.

## Introduction

### Rationale for the clinical research question

According to the United States’ Centers for Disease Control and Prevention (CDC), age-adjusted suicide rates in the US rose 39% from 2002 to 2022, with 49,476 Americans lost to suicide in 2022.^1^ Suicides are associated with a range of demographic and clinical risk factors, including male sex, religious affiliation,^2^ rurality, access to lethal means, and – among the most consistent predictors – severe pain^3^. Severe pain is also associated with suicide antecedents, including suicide ideation, plan, and attempt. The close association between pain and suicide has reignited concern in the United States, as news and scientific reports have shown increased rates of suicidal events among patients following reductions in daily prescribed opioid medications.^4–7^ This protocol paper describes the rationale for a research study that examines suicides occurring in the US among persons with a history of pain who have undergone reduction or taper of prescribed opioid medication. Guided by the Social-Ecological Model, this protocol paper also describes the development and application of an approach to psychological autopsy adapted to understanding suicide in persons with pain, with special focus on methods of recruitment in the absence of a master database of family or friends bereaved by suicide loss.

In the US, daily prescription opioid taper and discontinuation have emerged as statistical correlates of suicidal events,^6–8^ drawing public attention in the wake of efforts to remediate a US crisis of overdose, opioid misuse, and opioid use disorder.^9–11^ (Table 1). Prescribers, often primary care or pain physicians, reduce opioid doses for many reasons. They may articulate a concern for safety,^12^ or respond to policy pressure,^13, 14^ or – increasingly – find themselves obligated to reduce or stop opioids when pharmacies decline to fill opioid prescriptions as a result of perceived liability or opioid litigation settlement terms.^15, 16^ Also, when patients on long-term opioids lose a prescriber, other clinicians may be unwilling to assume responsibility for their care, and/or the patients’ opioid prescription.^17, 18^

**Table 1.**
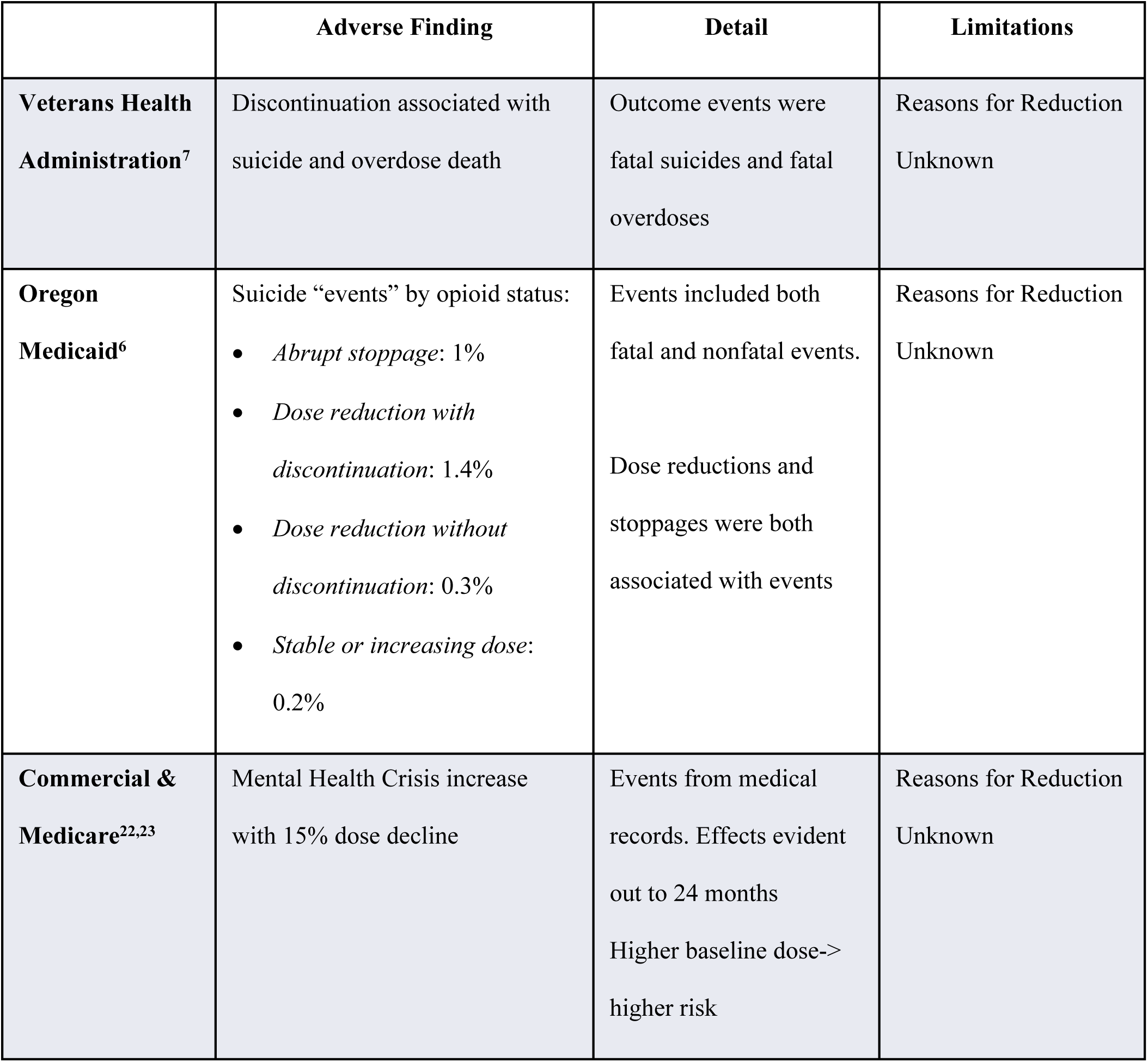
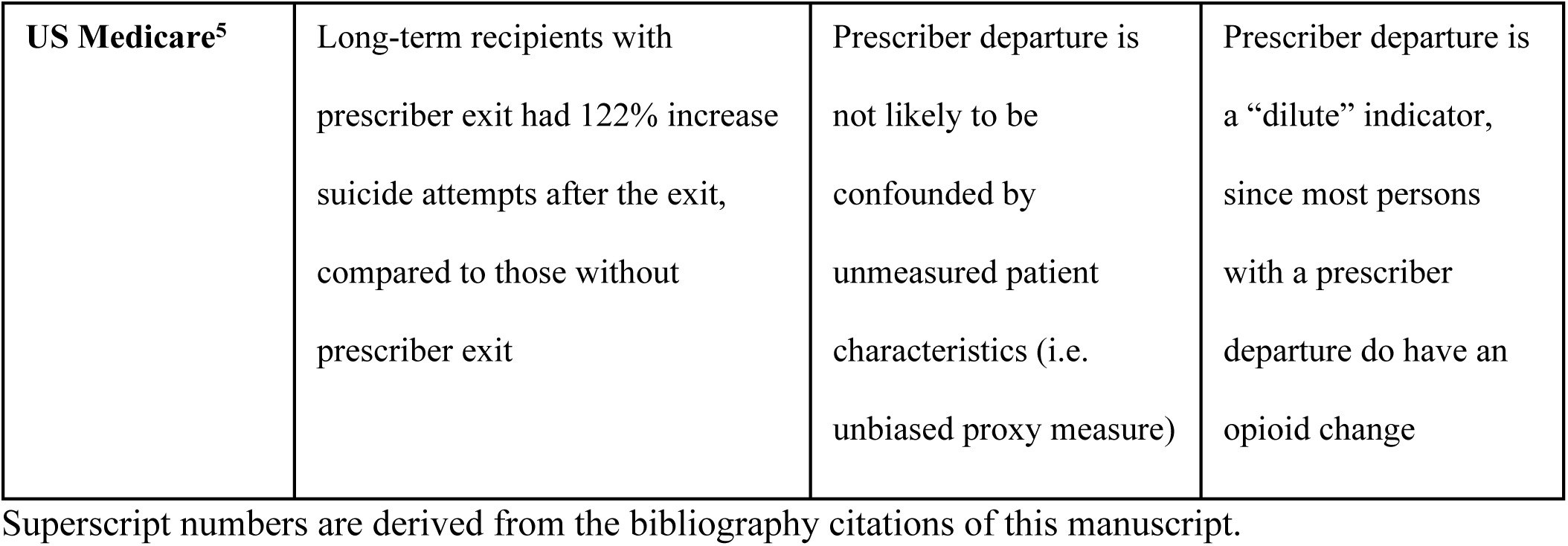
Selected Indicators of an Association between Prescription Opioid Reduction or Stoppage and Suicide-Related Events.

In addition to suicide and suicidal ideation, other adverse events resulting from abrupt opioid taper or cessation include mental health crises, overdose, and overdose deaths, thus prompting statements from the United States Food and Drug Administration (FDA),^19^ the CDC,^20^ and the US Department of Health and Human Services (HHS).^21^ All emphasize avoidance of rapid taper as central to avoiding a suicide outcome. For example, the FDA statement identifies only “sudden discontinuation” as a suicide risk, while the HHS report says, “Risks of rapid tapering or sudden discontinuation of opioids in physically dependent patients include acute withdrawal symptoms, exacerbation of pain, serious psychological distress, and thoughts of suicide,” while leaving ambiguous whether such risks apply to tapers that are not rapid.^21^ The shared emphasis on the rate of taper as determinative of risk is not wholly consistent with evidence, which suggests that taper rates are not likely to be the sole determinant of suicide risk.^6, 22, 23^

For example, a national insurance database study reporting on risk of mental health crisis (including suicide) found the risks were not restricted to persons rapidly tapered.^23^ Another paper reported elevated risk of mental health crises 12 months and later after opioid reduction.^22^ A smaller retrospective study of 2014-2015 Oregon’s Medicaid patients taking doses at higher than the equivalent of 50 milligrams of morphine per day found both suicide risk and other risk for opioid-related harm to be higher among patients who underwent opioid dose reduction prior to discontinuation, compared to patients who discontinued suddenly, even though those subject to initial reductions had been on lower doses.^6^ Such data are hard to square with attribution of suicide risk primarily to acute opioid withdrawal, although other factors of the taper process could matter (and were not reported in these studies): duration of taper, whether it was consensual, whether the outcomes were closely monitored, and whether mental health care was offered.

Although retrospective health system database studies do hint at some kind of risk, quantitative analyses of prescription and diagnostic records are not fully adequate to the task of explaining why, when, and how a person dies by suicide. Administrative and diagnostic codes recorded by health systems are assigned by medical staff who must check boxes to close out a visit and who are pressed for time. They are not life historians. The resultant codes say little about a person’s life history, pain experience, social relationships, or indeed most of the matters central to suicide scholarship, including perceived burdensomeness, a thwarted sense of belonging, social isolation, or overall mental anguish and suffering, regardless of whether it aligns with a mental health diagnosis (termed “psychache”).^24^

Detailed investigation of individual suicides should, ideally, consider all these factors, treating suicide for what it is: a complex event that emerges from a constellation of risk and resilience factors in a particular social and community context. In this study, we draw on the Social-Ecological Model, which considers suicide as emerging from multiple levels: society, community, organization, interpersonal, and individual (Fig 1).^25^

**Fig 1.**
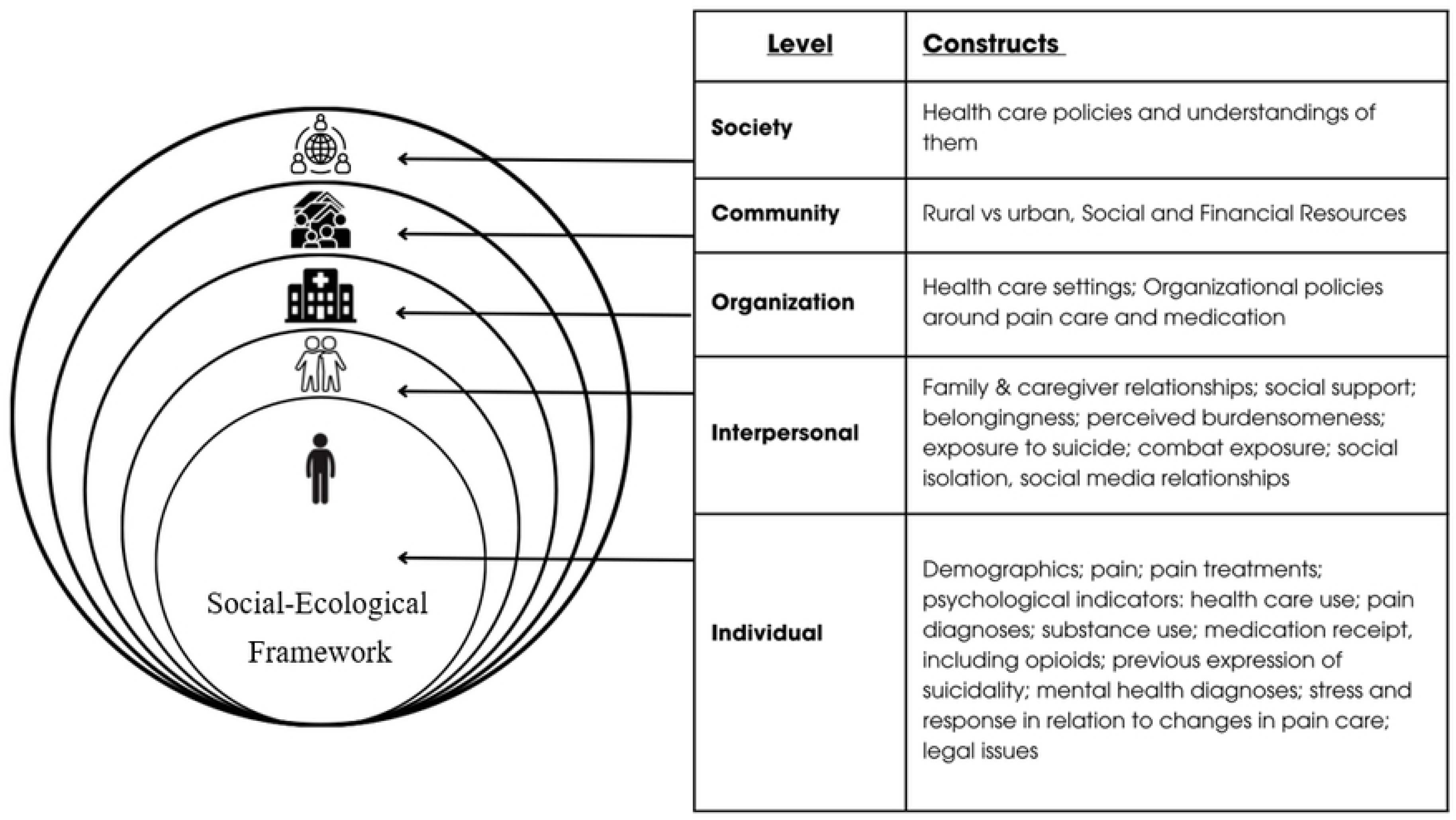
Social-Ecological Framework. Graphic adapted from Kok et al.^42^

### Rationale for an innovative methodology

The study Clinical Context of Suicide Following Opioid Transitions (CSI:OPIOIDs) seeks to examine suicides in a situation that has drawn public concern and speculation: those occurring after a prescription opioid dose reduction in patients with pain, including Veterans and non-Veterans.

Obtaining data about these suicides requires a substantial modification to traditional “psychological autopsy,” to our knowledge first described in 1957.^26, 27^ From inception, psychological autopsies combined interviews with the bereaved, and review of other data sources, as available, usually to develop quantitative comparative data.^28–31^ However, to our knowledge, there are no prior psychological autopsy studies in which recruitment was predicated on an antemortem health care event, a restriction presenting a challenge for recruitment, compared to traditional psychological autopsy research, where investigators recruit via outreach to a registry of all suicides for a jurisdiction.^32, 33^

Recruitment based on antemortem health care events presents legal and ethical barriers, especially in the US, where medical records and vital statistics systems are not integrated. Suicide attempt may appear in medical records,^22^ but suicide rarely does.^34^ Linking health system records to a national cause-of-death database for purposes of research recruitment is problematic. “Follow-back” from the US National Death Index data to bereaved family members of health care providers is usually prohibited owing to risk of emotional harm.^35^ For this reason, the present study makes a direct appeal to the public, aiming for in-depth qualitative rather than quantitative insight.

Public messaging outreach, however, must be calibrated to avoid conflict with social media platforms where terms such as “suicide” and “opioids” are restricted. Also, messages must be calibrated to substantial health system distrust among US communities with pain,^36^ many of whom have faced stigma^37^ or loss of care.^17^ Finally, a campaign mentioning suicide and opioid reduction could draw respondents who disproportionately attribute suicide solely to prescription change. This challenge may be less severe for a qualitative as opposed to quantitative study. In qualitative research, trustworthy knowledge can be developed when the methods of interview, record review, and interpretation are planned to assure that results are credible, confirmable, dependable, and transferrable.^38, 39^

This paper summarizes the protocol for the CSI:OPIOIDs study. This work began as a pilot recruitment demonstration in 2020. It was expanded into a full research project after receipt of funding by the US Department of Veterans Affairs in 2023. The study’s purpose is to characterize the patient and clinical context factors associated with suicide among Veterans and non-Veterans who have died by suicide in the context of opioid transition (stoppage or reduction), and to qualitatively compare the psychological autopsy findings between the Veterans and non-Veterans.

## Methods

### Conceptual framework

This study considers suicide as a complex multidetermined event, applying the Social-Ecological Model for suicide (Fig 1).^25^

The Social-Ecological Model for suicide seeks factors relevant to suicide at multiple levels: society, community, organization, interpersonal, and individual.^40^ Some opioid reductions result in suicidal crises, including lethal ones,^5, 6^ but others do not. An opioid medication change is likely to play out differently for people who differ in life history, support networks, and other factors. Careful exposition of the framework elements, in the life of each person who has died, could anchor future efforts to prevent such tragedies.^41^ Figure 1 shows relevant variables that can be identified from medical records and interviews with suicide loss survivors.^42^

The graphic of Figure 1 emphasizes domains we believe are likely important to pain and its care at the societal, community, organizational, interpersonal, and individual levels. It includes issues reported from observations of opioid prescribing reduction in the US, including changes in access to care, stigma, conflicts with pharmacies, and disability.^18, 36, 43–45^ The figure also incorporates aspects of the Interpersonal (or “interpersonal-psychological”) Theory of Suicide,^24^ which posits that both desire for and capability to die by suicide are important.^46–48^ That theory emphasizes both an intractably thwarted sense of belonging and the sense that one is a permanent burden to others.^46^

### Research design overview

The overall design of the study’s recruitment and data flow is summarized in Figure 2.

**Fig 2.**
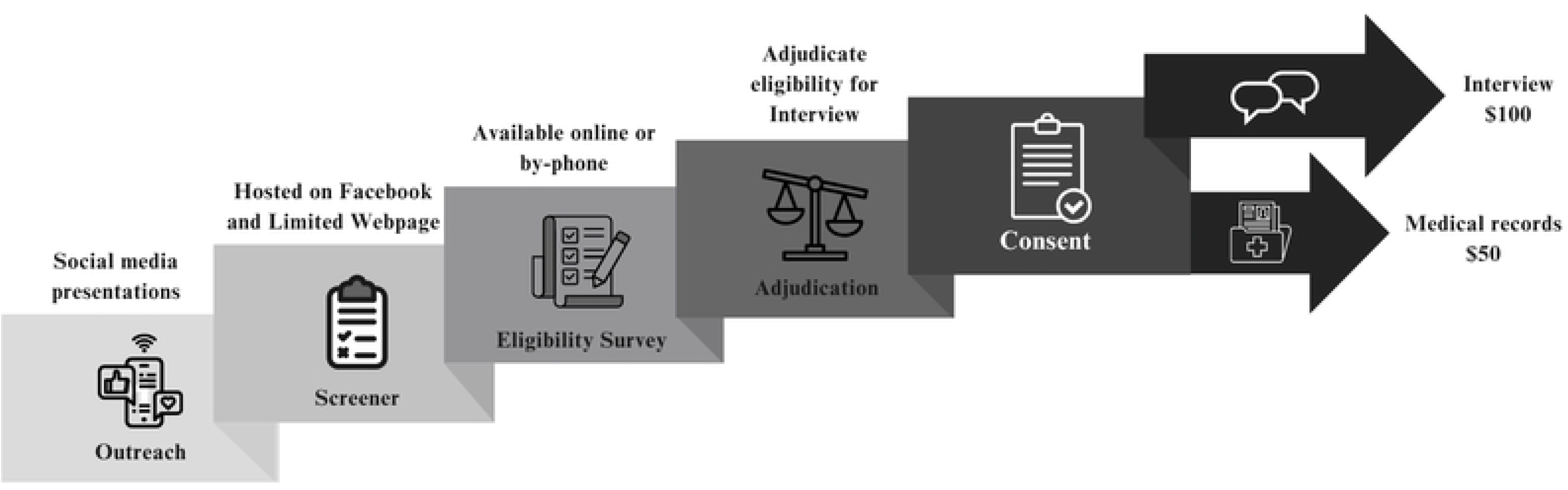
Recruitment and Data Flow.

This study recruits bereaved survivors of persons who are thought to have died by suicide in the context of an opioid dose reduction from within the US. Potential participants are recruited via social media marketing, and those interested are asked to respond to an online 46-item eligibility survey. Survey responses are adjudicated by the study team (see **Eligibility**). Eligible participants are invited to participate in an online-based semi-structured psychological autopsy interview. After in-depth interviews, both the interviewer and notetaker rate the interview for quality and depth of knowledge. A templated matrix analysis is used to analyze qualitative data.^49^

The proposed number of interviews targeted is 100-115, or until data saturation is reached. This will be determined by thematic saturation in the absence of new themes emerging in subsequent interviews.

In some cases, medical records and other relevant documents are obtained when the interviewee has authorized access. Interpretation of case histories is systematic and qualitative, seeking to identify common themes, such as early life risk factors, transitions in care, and crisis events that may be amenable to future program intervention. Once data from all sources have been collected and summarized, data will be integrated into a single case report for each decedent which includes the templated matrix.

### Psychological autopsy methodological considerations

Psychological autopsy studies present recruitment challenges that pertain to generalizability and validity.^28, 29, 50^ These challenges take a different form in a qualitative rather than quantitative study.

### General considerations related to recruitment

Traditional psychological autopsy research differs from the present study in that such traditional research seeks all suicides from a jurisdiction, with investigators recruiting in collaboration with medical examiners possessing access to a jurisdictional registry and offering quantitative summation of risk factors.^26, 32, 51^ Such studies include a quantitative comparison between decedents, and potentially—controls who did not die—or died by other means.^30, 52^

This approach does not fit with efforts to study deaths by suicide after a prescription opioid reduction or stoppage, at least within a US health care context. The hypothetical “list” of relevant decedents does not exist, and the number of deaths in any single jurisdiction would be small. Medical examiners could not produce such a list as they often lack access to antemortem health records.

Considering these challenges, this project team implemented direct-to-public outreach as the primary recruitment strategy, including advertising and public presentations.

### Overview of personnel

The team structure is summarized in Figure 3 and briefly summarized here, given the breadth of experience and expertise required. Core investigators include faculty with research footholds in primary care, pain care, health service delivery, medical anthropology, health services research, implementation science, substance use disorders, care of vulnerable populations, and experience in statistical and qualitative methods. All interviewers obtained certificates in psychological autopsy through training with the American Association of Suicidology. Advisors with Lived Experience includes individuals with lived experience of suicide loss, pain and addiction, several paid as advisors and one as a paid employee. Consultants include experts in suicide, Veteran families, opioid issues, pain care, and postvention response for family stabilization due to suicide bereavement. Veterans Health Administration partner offices include the VA Office of Suicide Prevention, and the VA’s National Pain Management, Opioid Safety, and Prescription Drug Monitoring Program. The study also relies on contracts for social media marketing, interview transcription, assistance in management of the study’s eligibility survey, and procurement of medical records, when possible.

**Fig. 3.**
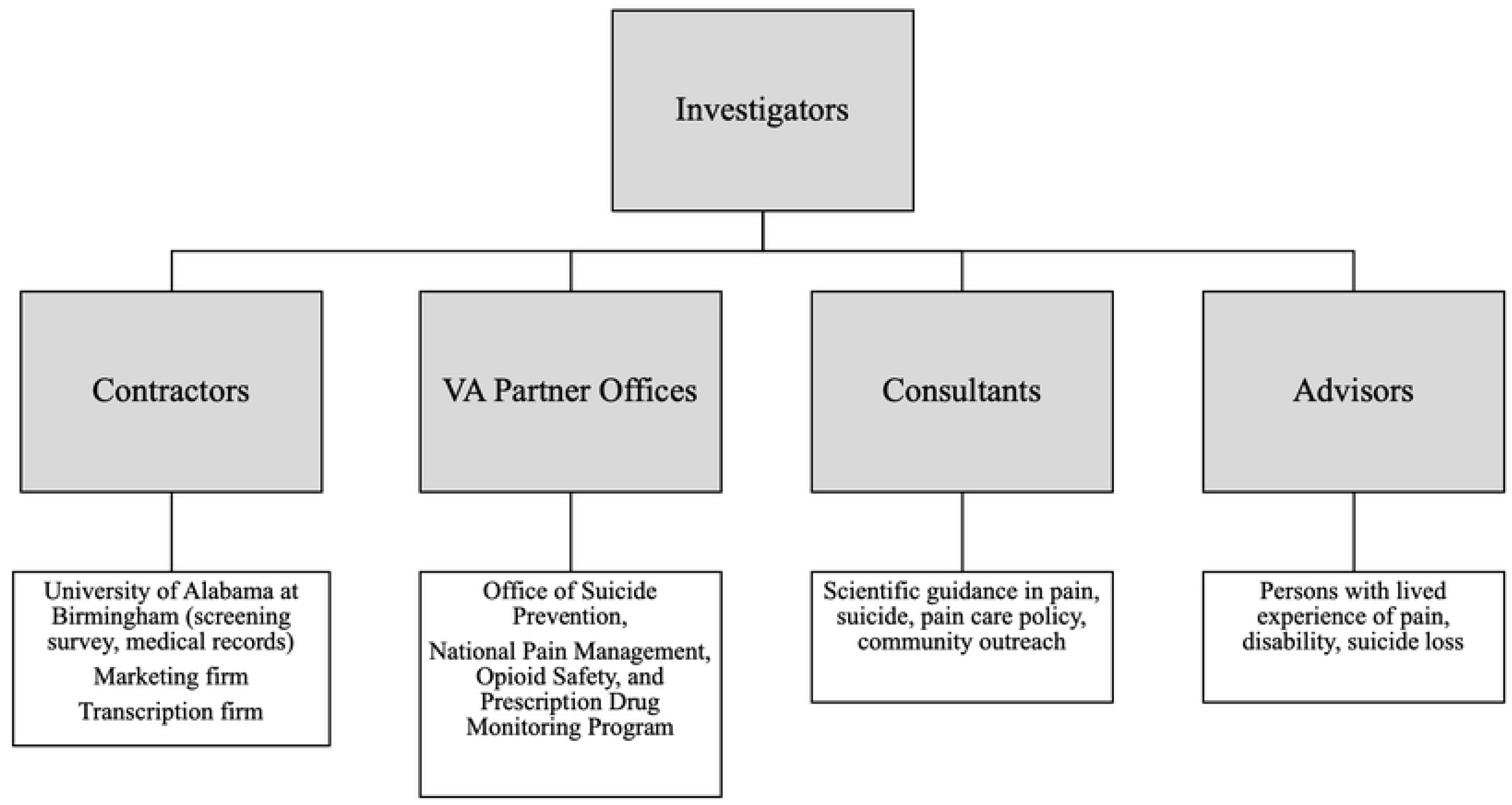
Team Structure.

### Ethical oversight and privacy

The study is subject to oversight by the Veterans Health Administration Central Institutional Review Board (IRB), and the IRB of the University of Alabama at Birmingham (UAB). Ethics authority for matters occurring under university auspices (recruitment, primarily) lies with UAB, as does the management of UAB-accrued recruitment data. Conversely, interview conduct and management of the resulting interview data, lie with the Veterans Health Administration. Study data are subject to privacy protections applicable to human subjects’ data through a Certificate of Confidentiality from the National Institutes of Health. Study team members who conduct interviews and take part in analyses have IRB approval to view identifiable participant information.

### Justification for the included population

Eligible persons are those who lost a family member or close friend with pain, to suicide after an opioid pain medication change. The persons who died by suicide are termed “decedents.” After some prescribers who lost patients approached the study to request participation, the IRB-approved protocol was modified to permit such prescribers/clinicians to participate. Adjudication of an eligibility survey is required to assess which potential participants appear to be reporting a suicide with a sufficient confidence that the death was by suicide, in a person with pain, and with sufficient confidence that it transpired after a reduction or stoppage in pain medication. This is reviewed under **Eligibility** below.

Restriction based on recency of death was considered, given the informational value of fresh memories. However, given the absence of prior studies of this nature and the risk of under-recruitment, the timespan for eligible deaths was kept expansive, back to those thought to have happened after 2012, the year that opioid prescriptions peaked in the US.^53^ The variability in time elapsed and in the types of respondents, led us to add interviewers’ assessments of trustworthiness of data into the coding process for each interview as shown below in **Oral interview with survivor.**

### Recruitment

The primary recruitment mode involves advertising and distribution (paid or unpaid) through social media, newsletters, and similar fora as introduced under **Psychological autopsy methodological considerations** above. The recruitment process, predicated on collaborative work between VA-based and university-based research teams, includes components summarized in Figure 2.

### Public outreach in pilot phase

A pilot version of CSI:OPIOIDs, prior to receipt of external funding, initiated public recruitment in November 2020. That version was advertised primarily by press release and social media graphics created by members of the investigative team and then expanded through a small contract to a research recruitment firm. Sixty-eight respondents completed the full pilot survey and expressed their willingness to be contacted as of July 2023. The pilot study relied on the same preliminary screening survey as the ongoing main study. Once major grant funding was underway, the study team reached out to those qualifying under eligibility criteria for interview.

### Advertising campaign

A public advertising campaign with a designated, contracted firm with appropriate expertise in social media campaigns for research facilitates public outreach for the recruitment of study participants. Advertisement images vary with respect to demographic characteristics, including race, gender, and Veteran status, and are disseminated through paid media and social media, including Facebook.

### Targeted outreach

Partnered outreach with key organizational allies was not required as part of the study. However, members of the investigative team prioritize opportunities to speak to families as well as advocacy and scientific groups. Any organization or individual with access to individuals who are bereaved by loss of someone to suicide can freely distribute information provided by the study, including single-page handouts and links to social media.

### Eligibility

The decision concerning who to invite for interview depends on an **eligibility survey** (Supplement 1), and adjudication of responses (Supplement 2). This survey is hosted by the University of Alabama at Birmingham Heersink School of Medicine on a secure REDCap server. An individual may navigate to the recruitment survey directly online. Alternatively, they may first encounter a short preliminary screening survey (e.g. on Facebook) that invites proceeding to the eligibility survey.

The eligibility survey can be completed online or by phone. It has varied in length from 44 to 52 items from inception during the study’s pilot phase to the time of this writing. Although 52 items at inception, the full screener was adjusted to 46 in mid-2024, primarily by removing some detailed and, at times, partly redundant items.

On the eligibility survey, initial items query the participant’s age (19 years or older is required under the laws of Alabama), whether they are in the United States, the decedent’s last location, and preference for methods of contact. Crucially, the very first item asks, “*Do you believe that someone close to you died by suicide after a change in pain medication?*” The word “*after*” was chosen to avoid direct assignment of causation. Persons responding affirmatively and meeting criteria for age and location are invited to proceed. Those responding negatively are offered the opportunity to provide contact information for future studies.

The eligibility survey asks the respondent to register their level of certainty or uncertainty both regarding (a) the nature of the death being a suicide and (b) the type of medication change (**Fig 4 and Sup 1**).

**Fig 4.**
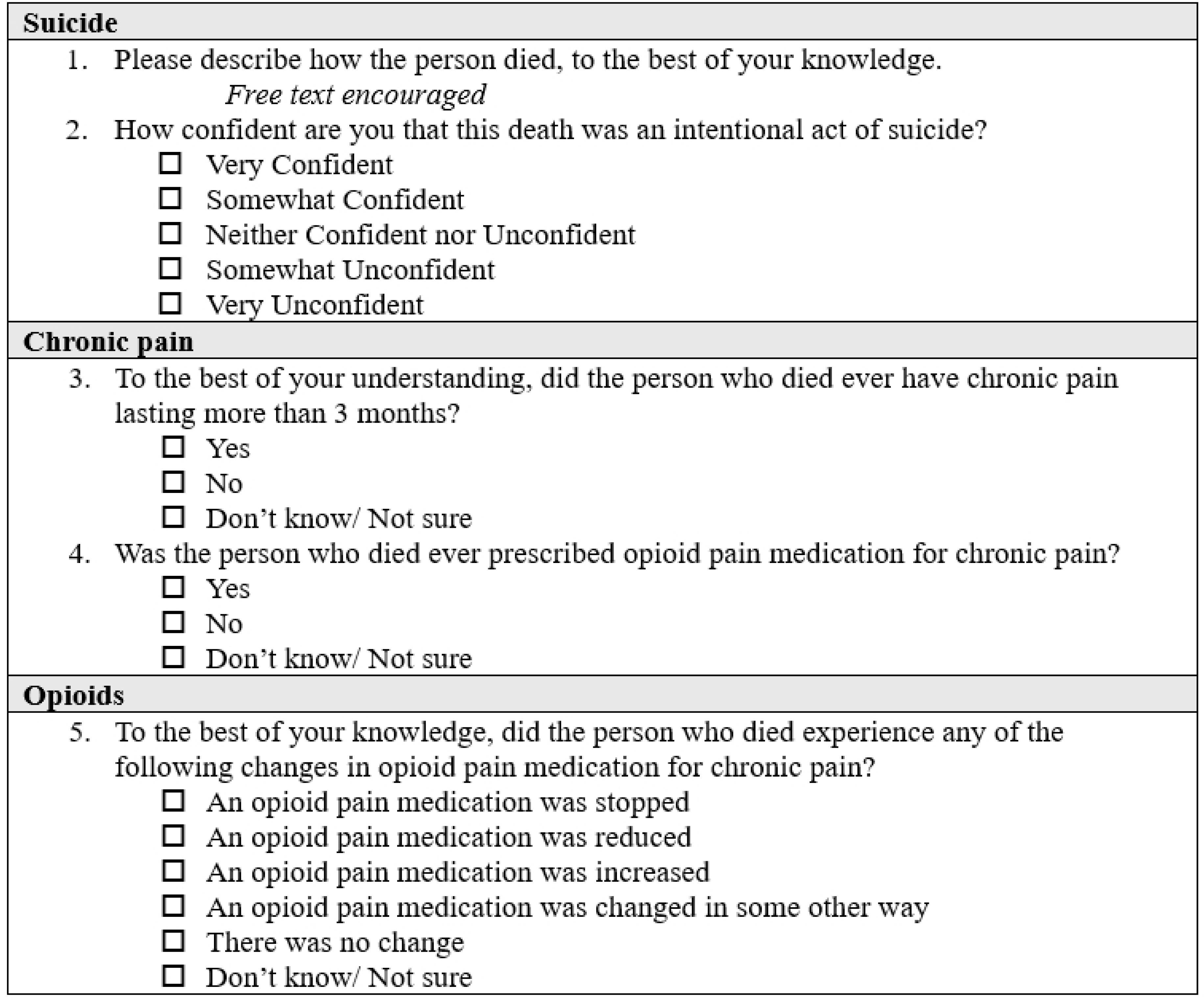
Eligibility Survey Items Concerning Suicide, Pain, and Opioid Reduction.

For the adjudication of a likely prescription opioid reduction, eligibility is based on the response to the prescription opioid reduction survey item. Respondents are allowed to check multiple responses (for example “an opioid pain medication was reduced” and “an opioid pain medication was changed in some other way”) to account for the possibility that informants may observe changes in multiple distinct opioid pain relief medications without possessing expertise on potency sufficient to judge which medicine was the more potent or less potent, and so on.

Response options permit a statement of uncertainty, and free-text optional responses are checked as part of the adjudication process. For the adjudication of likely suicide, a similar logic is applied.

In July 2024, the following question was added to the detailed screener: “To the best of your knowledge, did the person ever have chronic pain lasting more than 3 months?” The situation leading to this change consisted of 4 interviews where the interviewee acknowledged uncertainty as to whether the decedent ever had long-term pain. With this adjudication, a “yes” response was further required to qualify for the invitation.

The overall design of the study’s recruitment and data flow is summarized in the figure titled “**Recruitment and Data Flow**” under Figure 2. This includes:

(A) public advertising and outreach, leading to screeners (detailed and, in some instances, preliminary)
(B) written informed consent (via postal mail or VA DocuSign)
(C) medical records collection (when possible)
(D) interviews with survivors
(E) analyses/synthesis

This overall plan is carried out through collaboration with UAB, responsible primarily for recruitment and soliciting non-VA medical records, and VA. At the time of submission, recruitment advertising was continuing.

### Distress and safety

Psychological autopsy interviews are sensitive in nature, and they may cause temporary emotional upset, although published studies indicate they do not cause major or lasting distress, and a majority of participants find them helpful.^54^ The study team has safety protocols for response to persons indicating distress. During the first 39 interviews, there arose no occasion requiring such a response.

Somewhat unique to this study, the recruitment process entails public outreach likely to be seen by US patients with long-term pain and opioid receipt, who face an ongoing loss of care.

In the US, living persons with pain and a history of prescription opioid receipt are increasingly subject to abrupt prescription opioid taper,^55^ and rejection from medical practice by clinicians unwilling to receive them.^17^ The investigative team judged it likely that some individuals would contact the research team in desperate efforts to find care. Considering our inability to assume direct care responsibilities, the following steps were taken. First, the principal investigator, a physician, posted an independent, non-study-affiliated guide for families and patients facing unwanted opioid medication reduction. Second, a study website includes stories of work by members of the team to indicate non-research efforts to improve care through advocacy. Finally, individualized responses to distress emails clarify study boundaries while directing individuals to appropriate resources, including the aforementioned family guide.

Additional safety protocols were crafted to govern interactions between the recruitment team at UAB (UAB’s Recruitment and Retention Shared Facility) and members of the public who interact with the study through its survey screening process, either by email, online survey, or telephone (Supplement 3). As a general matter, distress may include declarations of ongoing emotional crisis (termed “Category 1”) and simple requests for assistance (termed “Category 2”). The Category 1 requests include efforts to achieve a “warm hand-off” to a crisis support line if the individual is in live contact on the telephone, or response involving rapid contact to a clinically trained individual if not. The Category 2 situations have primarily involved requests for advice in relation to ongoing changes to pain care, which are usually handled with both a personalized email and reference to the published guidance by the principal investigator.

### Oral interview with survivor

The CSI:OPIOIDs detailed interview guide, shown in Supplement 4, is based on the outline of the interview guide used by the Veterans Administration Behavioral Health Autopsy Program,^56^ subject to modification based on study interests and components of the study’s Social-Ecological Model for suicide, as well as the logistics of the interview. The interview guide development and modification (January-June 2023) included review and input by experts in suicide, and a range of disciplinary experts including health services research, pain, medical anthropology, public health, and medicine. The interview guide was pilot tested with a study advisor in April 2023, an individual with chronic pain who was a close friend to someone who they believed had died by suicide. Based on the pilot run, it was further modified, and further adjustments were made during the first five data collection interviews.

The interview combines qualitative and closed-ended survey items. Its contents are summarized in Table 2. Interviews are recorded and saved through VA’s Microsoft Teams, where the participant may either participate through the application or by telephone.

**Table 2.**
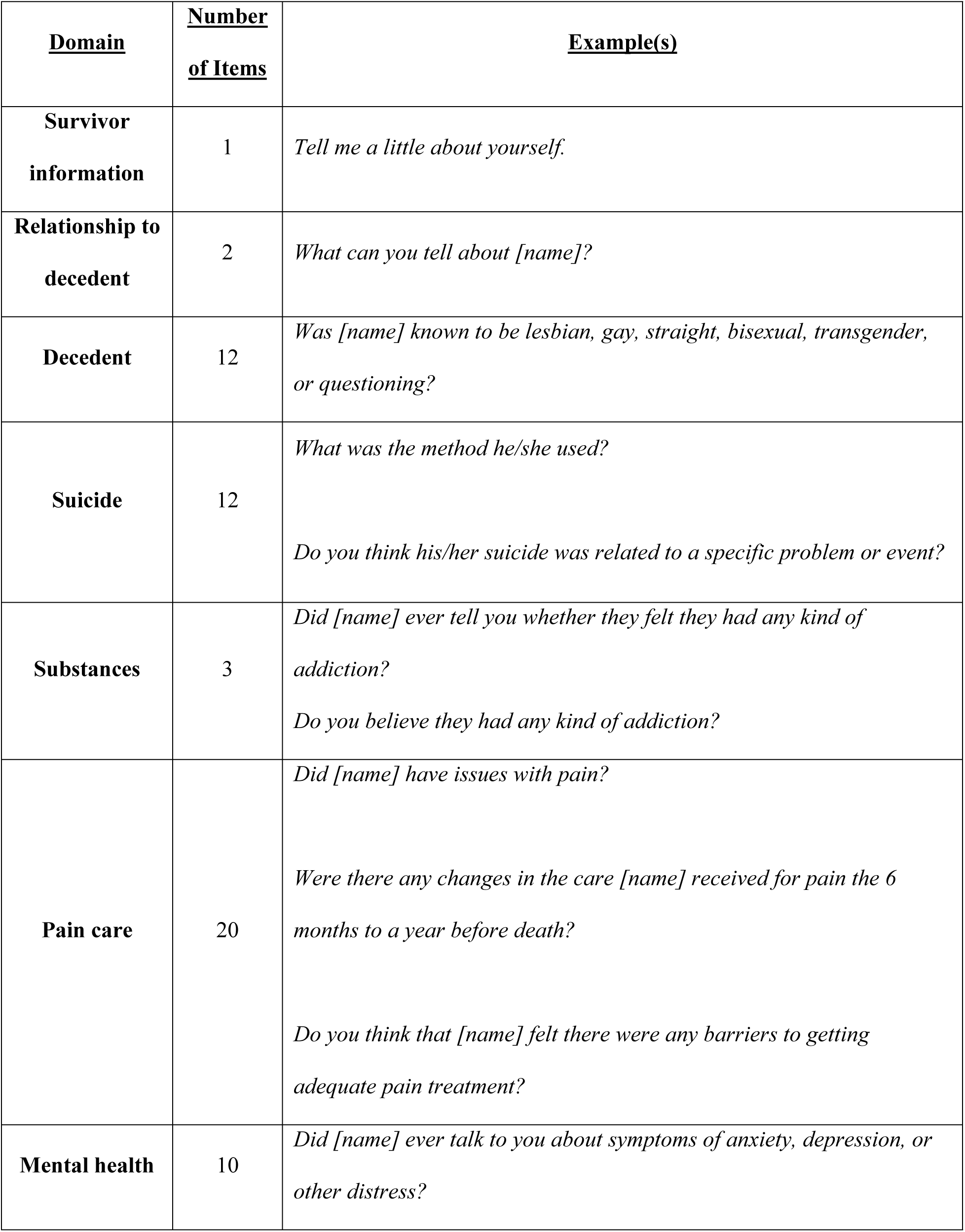
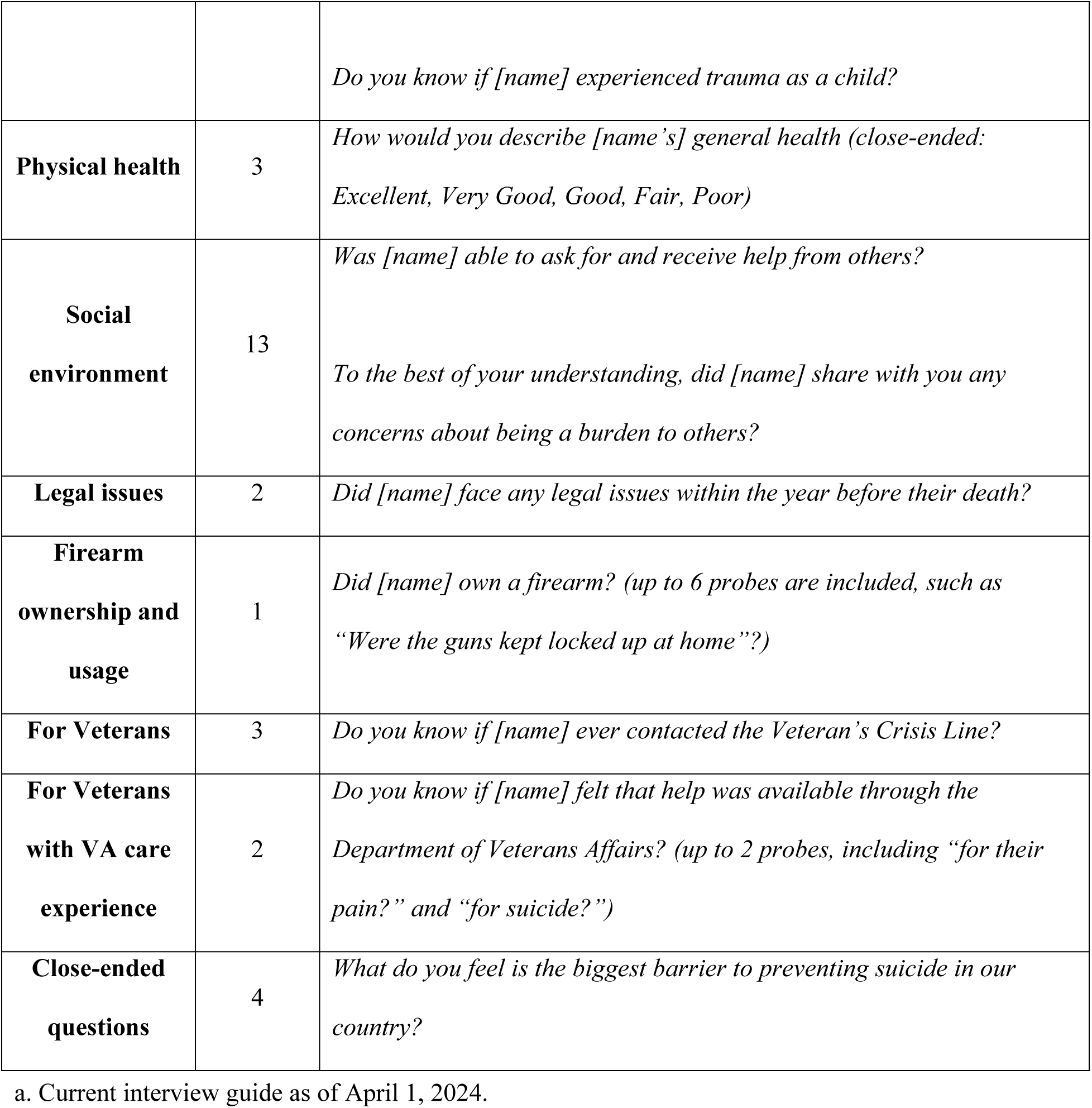
Interview Guide Contents Summary.^a^.

| **<u>Domain</u>** | **<u>Number</u>**<br>**<u>of Items</u>** | **<u>Example(s)</u>** |
| --- | --- | --- |
| **Survivor**<br>**information** | 1 | *Tell me a little about yourself.* |
| **Relationship to**<br>**decedent** | 2 | *What can you tell about [name]?* |
| **Decedent** | 12 | *Was [name] known to be lesbian, gay, straight, bisexual, transgender,*<br>*or questioning?* |
| **Suicide** | 12 | *What was the method he/she used?*<br>*Do you think his/her suicide was related to a specific problem or event?* |
| **Substances** | 3 | *Did [name] ever tell you whether they felt they had any kind of*<br>*addiction?*<br>*Do you believe they had any kind of addiction?* |
| **Pain care** | 20 | *Did [name] have issues with pain?*<br>*Were there any changes in the care [name] received for pain the 6*<br>*months to a year before death?*<br>*Do you think that [name] felt there were any barriers to getting*<br>*adequate pain treatment?* |
| **Mental health** | 10 | *Did [name] ever talk to you about symptoms of anxiety, depression, or*<br>*other distress?* |
|  |  | *Do you know if [name] experienced trauma as a child?* |
| **Physical health** | 3 | *How would you describe [name's] general health (close-ended: Excellent, Very Good, Good, Fair, Poor)* |
| **Social environment** | 13 | *Was [name] able to ask for and receive help from others?*<br>*To the best of your understanding, did [name] share with you any concerns about being a burden to others?* |
| **Legal issues** | 2 | *Did [name] face any legal issues within the year before their death?* |
| **Firearm ownership and usage** | 1 | *Did [name] own a firearm? (up to 6 probes are included, such as "Were the guns kept locked up at home"?)* |
| **For Veterans** | 3 | *Do you know if [name] ever contacted the Veteran's Crisis Line?* |
| **For Veterans with VA care experience** | 2 | *Do you know if [name] felt that help was available through the Department of Veterans Affairs? (up to 2 probes, including "for their pain?" and "for suicide?" )* |
| **Close-ended questions** | 4 | *What do you feel is the biggest barrier to preventing suicide in our country?* |

a. Current interview guide as of April 1, 2024.

After the first 4 interviews, we encountered participants with a wide range of knowledge of the decedent. Therefore, we developed ratings for credibility of the interviewee, which are assigned by both interviewer and the notetaker. Both assign a 1-5 score based on their perception (5 represents a respondent who is thought to be highly knowledgeable, and 1 is the lowest possible level of apparent knowledge). Scores are not used to formally exclude interview findings but can inform interpretation.

Participant interviews began in August 2023. It is anticipated that recruitment will be completed by January 2026 with data (interviews with each participant) by March 2026 and analyses by August 2026.

### Medical records acquisition

Medical records are only obtainable if the study respondent holds authority to request them, and the health care agencies prove responsive. A professional subunit of UAB School of Public Health experienced in records procurement is contracted to work with informants to obtain records, scan them, and assist in conveyance to the VA study team. When care has transpired within VA itself, VA research staff can obtain them internally, if the informant for the study has authority to release them.

### Medical records review

Obtainable records are expected to be irregular. That is, they will be from various clinics, of varying relevance to the pain care history, with varying time covered. A detailed review of every page is unlikely to be informative. A template for records review focuses primarily on aspects of care that are likely to be determinable by a trained clinician within a few hours of review, considering that outpatient visit notes do not routinely offer systematic pharmacy records. The research team, which included several clinicians familiar with strengths and weaknesses of how pain care is documented, developed a set of record-derivable care indicators. Those indicators and the template for review are shown in Table 3, “Medical Record Review Guide”.

**Table 3.**
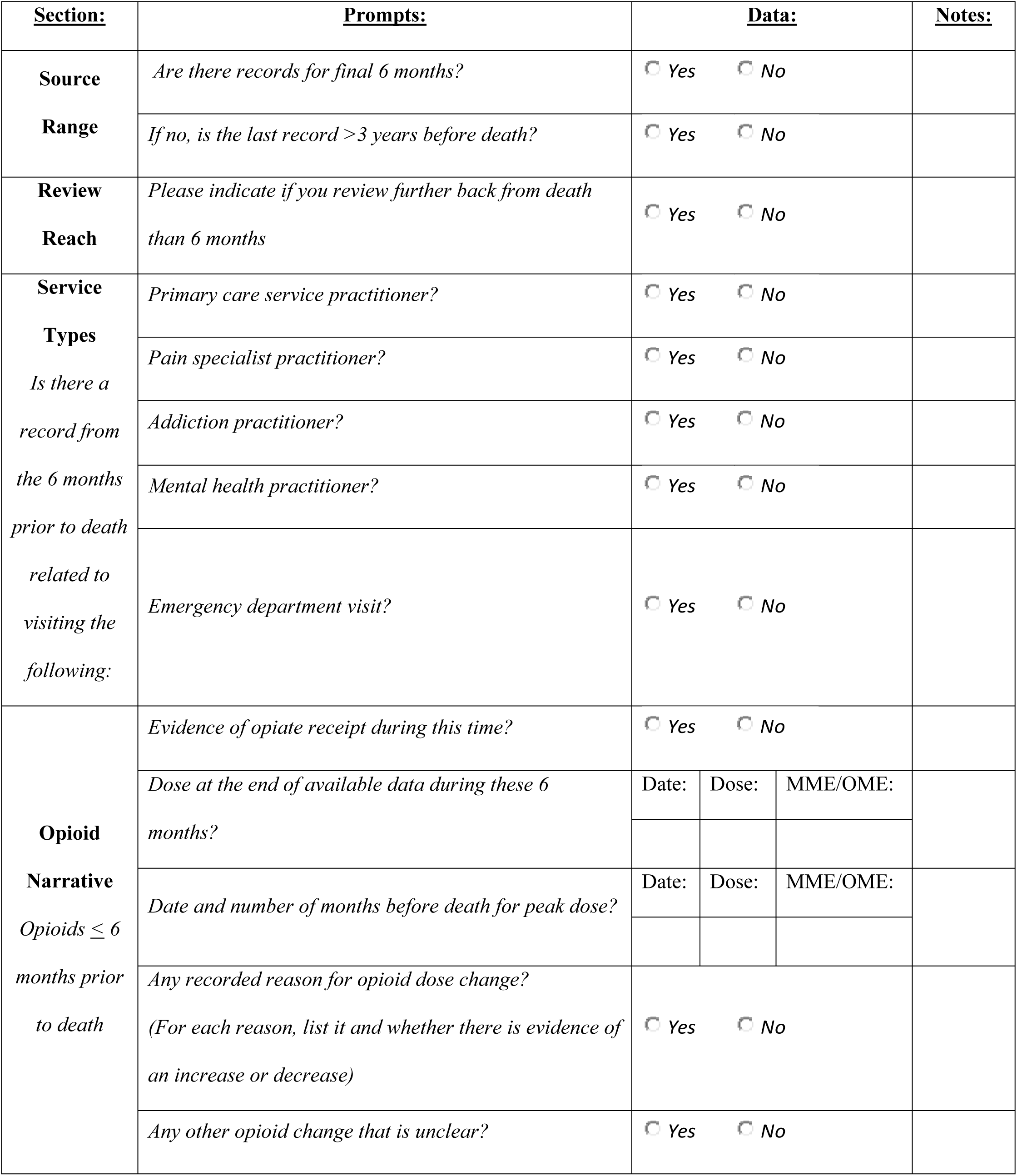
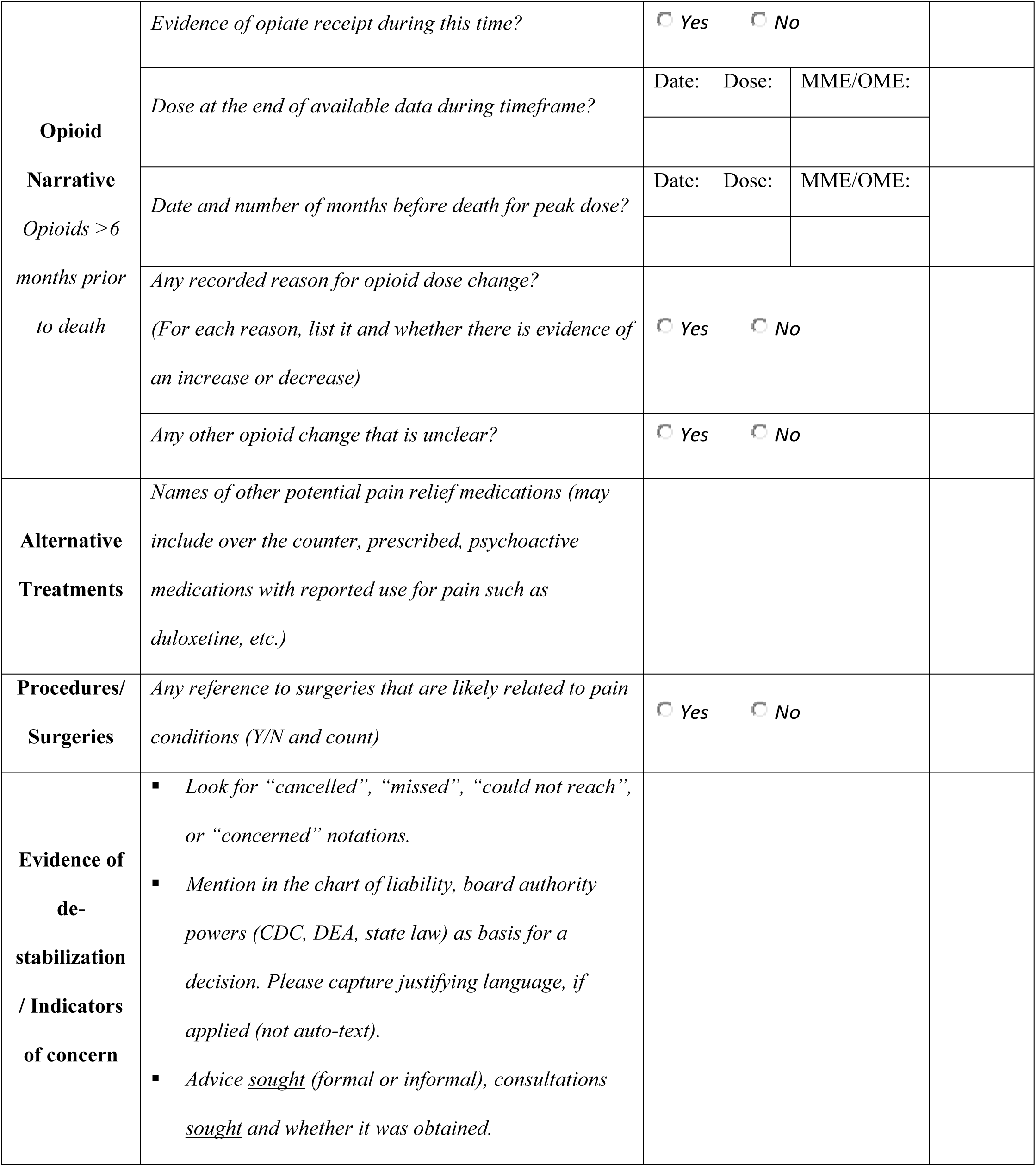
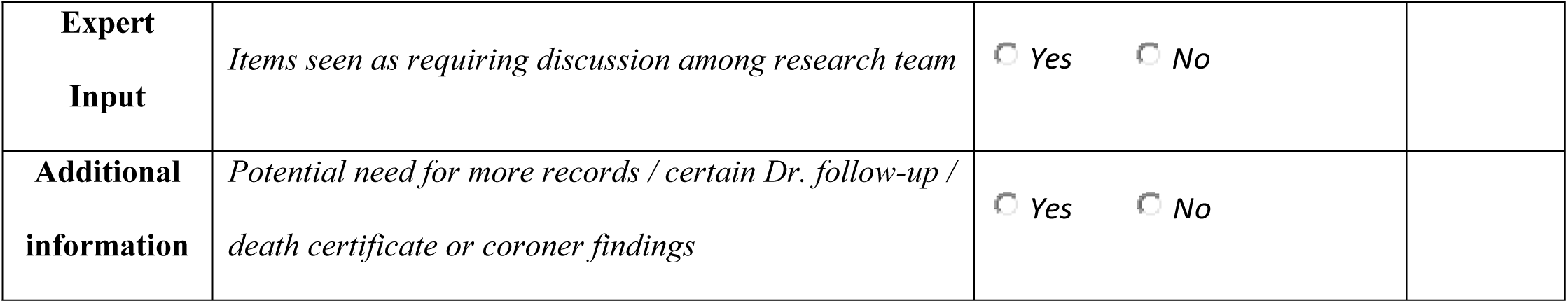
Medical Record Review Guide.

### Other sources of data

The study design incorporates the potential triangulation of information from the interview with what may be available based on suicide notes, correspondence with the study participant, or publicly accessible social media. When provided by the informant, they are saved alongside the interview transcript and incorporated into the coding process.

### Analysis of interview content

Transcribed interviews are reviewed and subject to review by a multidisciplinary team of 2 to 5 individuals by consensus. The approach tags and codes standardized information for each source interviewed. The information conforms to topical areas queried within the interview, and topical concerns that emerge during transcript review. This approach aligns with templated analysis.^57, 58^ However, multidisciplinary deliberation includes focused and open discussion and debate regarding apparent gaps in the participant’s knowledge. It includes efforts to sift and interpret the respondent’s utterances and interpretations through review of the relationship between the participant and the decedent.

### Quantitative analysis

The proposed comparison of reports of Veteran and non-Veteran decedents is likely to permit some quantitative comparisons for categorical and, regarding age, continuous variables. However, because the recruitment methods are opportunistic and purposeful, findings derived from quantitative analysis cannot be generalized to a hypothetical population.

### Ethics

Human subjects in a study are, by definition, **living** (**bereaved) survivors** (friends or family members) who seek to participate in the study. Ethical obligations, however, extend to persons who contact the study even if their preliminary survey responses do not qualify them for entry. Safety protocols laid out above apply for both groups.

### Confidentiality

All research studies in the US are subject to stringent confidentiality requirements. In the US legal context, additional protections are prudent to protect study participants from legal demands for production of research data in case of criminal investigation or civil litigation discovery. The latter situation arises when patients’ families initiate legal action based on a death, as has happened within the US.

A “Certificate of Confidentiality”, a US legal protection for participants, is conferred automatically only to studies funded directly by the National Institutes of Health. For studies funded by the US Department of Veterans Affairs, such certificates are not offered automatically but require a special request. Once we learned of participants with grave fears regarding the confidentiality of data, we requested and obtained NIH confidentiality certificates for both the VA and University-based components of this study.

## Discussion

This paper describes, to our knowledge, the first qualitative psychological autopsy study to examine, in depth, suicides after opioid medication reduction in persons with pain. Statistical reports on this matter ^5, 59–61^ are numerous and have spurred cautionary guidance from some agencies.^19, 20^ Those agency statements, with their emphasis on slow versus rapid taper, tend to imply that acute opioid withdrawal is the actionable and preventable cause of suicide in this population.

That public emphasis on preventing acute withdrawal, however, does not align with statistical evidence.^22^ It is at odds with contemporary conceptual understandings of suicide, which rarely attribute suicide to a specific physiological cause.^24, 62^ Notably, available retrospective studies correlating suicide or related crises with prescription opioid reduction report suicide events and mental health crises transpiring months to years after the medication change, well outside the period of acute withdrawal.^7, 22^ Pain itself may recur after medications are stopped. It is a recognized risk factor for suicide,^3^ but pain alone is not a sufficient explanation for death by suicide. Modern scholarship on suicide approaches this tragedy as the result of a collision of multiple factors in a person’s life, unfolding over time, in a particular narrative context.

Statistical profiles developed from standardized extracts of health care records have value. However, the diagnostic codes applied to US medical records are often inaccurate,^63^ and they are not designed to capture the full range of individual and social factors that could lead to a human being ending their own life.

This study protocol involves a qualitative adaptation of psychological autopsy, a method used in suicide research since at least the late 1950s.^26^ Our approach was, to some degree, shaped by recruitment constraints, as suicide cases had to be identified through direct public engagement rather than drawn from all deaths within a given catchment area. A statistically representative sample of **all** suicides linked to a unique premortem health care event is not possible under contemporary permissible uses of US-based health records and death records. In other words, there is no official master list of suicides that follow prescription opioid reduction in the US. Even if there were, contacting bereaved survivors randomly and unannounced, based on such a list, would invite serious ethical concerns.

Recruitment for this study is, as a result, unique. It depends on public messages, paid advertising, and an online eligibility survey. The resulting information will reflect the types of people who judge it meaningful or useful to discuss a tragic loss. It affords qualitative but not quantitative insight.

However, qualitative inquiry remains particularly helpful when trying to understand how a wide range of influences transpired in each person’s life to lead them toward taking their own lives. Qualitative health services research is often case-based. Such an approach contributes to theory-building, and it highlights potential gaps or unintended consequences in care that are not routinely identified or recorded in any other way.

The approach does take account of the inherent limitations of knowledge for the informant reporter. Interviewers are trained and skilled in qualitative interview and in psychological autopsy itself. The interview protocol is standardized. The individuals doing the coding include individuals with scholarly domain expertise (i.e. health systems, medical anthropology) and clinical expertise (treatment of pain, opioid use disorder). Their interaction aims at identifying credible interpretations and averting speculative conclusions.

## Conclusions

The ideal outcome for this study will be a detailed profile of common risk factors and event patterns related to suicides in persons with pain who undergo prescription opioid reduction. These events may include interruptions of health care, absence of care, breakdowns in clinical or social relationships, specific non-health crises occurring in the time prior to death, or patterns in the types of understandings or misunderstandings that transpire in an individual’s social environment prior to death by suicide. Some of these patterns may point to direct action that can be pursued in short order. Others will require systematic exploration with study advisors, including clinicians and people with pain and with suicide experts to identify future testable interventions.

## Data Availability

N/a

## Authors’ Contributions

**Stefan G. Kertesz**

Roles: Conceptualization, Funding acquisition, Investigation, Methodology, Project administration, Supervision, Writing-original draft, Writing-Review and editing

**Allyson L. Varley**

Roles: Conceptualization, Funding acquisition, Investigation, Methodology, Writing-original draft, Writing-Review and editing

**April E. Hoge**

Roles: Conceptualization, Funding acquisition, Investigation, Methodology, Project administration, Supervision, Writing-original draft, Writing-Review and editing

**Thomas E. Joiner**

Roles: Conceptualization, Funding acquisition, Methodology, Writing-Review and editing

**Beth Darnall**

Roles: Conceptualization, Methodology, Writing-Review and editing

**Kate Nicholson**

Roles: Conceptualization, Methodology, Writing-Review and editing

**Anne Fuqua**

Roles: Conceptualization, Investigation, Writing-Review and editing

**Mary F. Gilmore**

Roles: Investigation, Project administration, Writing-Review and editing

**Kevin R. Riggs**

Roles: Investigation, Methodology, Writing-Review and editing

**Stephanie Gamble**

Roles: Conceptualization, Methodology, Writing-Review and editing

**Adam J. Gordon**

Roles: Conceptualization, Funding acquisition, Investigation, Methodology, Supervision, Writing-Review and editing

**Yogesh Dwivedi**

Roles: Funding acquisition, Methodology, Writing-Review and editing

**Mark Flower**

Roles: Methodology, Writing-Review and editing

**Christin Veasley**

Roles: Funding acquisition, Methodology, Writing-Review and editing

**Carla Stumpf Patton**

Roles: Conceptualization, Funding acquisition, Methodology, Writing-Review and editing

**Ashley Leal**

Roles: Conceptualization, Funding acquisition, Methodology, Writing-Review and editing

**Dawn Gibson**

Roles: Conceptualization, Methodology, Writing-Review and editing

**Jim Elliott**

Roles: Conceptualization, Methodology, Writing-Review and editing

**Megan B McCullough**

Roles: Conceptualization, Funding acquisition, Investigation, Methodology, Supervision, Writing-Review and editing

## Disclaimers and Acknowledgments

Views expressed in this manuscript represent views of the authors and do not represent views or positions of the United States Department of Veterans Affairs. The authors acknowledge the kind assistance and advice of numerous advisors to the study, as well as the Director of Pain Programs for the VA, Friedhelm Sandbrink, and the VA Office of Suicide Prevention.

## Supporting information

**S1 Appendix. Eligibility Questionnaire.**

**S2 Appendix. Adjudication of Eligibility Survey.**

**S3 Appendix. Safety Protocol for Recruitment and Retention Shared Facility.**

**S4 Appendix. Detailed Interview Guide.**

